# Patients’ preference and time to finish injection for GnRH agonist and GnRH antagonist in Japanese prostate cancer patients

**DOI:** 10.1101/2023.08.25.23294461

**Authors:** Takashi Kawahara, Hiroji Uemura

## Abstract

Huggins et al. introduced surgical castration for prostate cancer treatment in 1941. Currently, androgen deprivation therapy (ADT) is key in treating advanced prostate cancer, especially in elderly patients. With the development of gonadotropin-releasing hormone (GnRH) medications, ADT transitioned from surgical to medical castration. From 1985 onwards, various GnRH agonists like Leuprorelin and Goserelin acetate have been used as ADT for prostate cancer. By 2012, a GnRH antagonist, degarelix, was introduced in Japan.

A study at Yokohama City University Medical Center analyzed 851 hormonal injections between August 2018 and February 2019. It assessed the duration from prescription to injection completion and the perceived physical and mental load on patients. Results showed degarelix injections took significantly longer than other treatments. Leuprorelin 22.5mg reduced hospital stays the most, largely due to its 6-month duration and kit formulation. The lengthy process for degarelix, which requires drug dilution and cooling at the injection site due to potential skin reactions, increases its administration time.

Concerning patient burden, leuprorelin 22.5mg ranked the lowest, with little variance from the oral form. A limitation of this study is the administration method of leuprorelin in Japan, which differs from other countries. In summary, the 6-month leuprorelin 22.5mg regimen minimizes hospital time, while degarelix tends to cause the most discomfort for patients.

## Main Manuscript

Huggins et al. firstly performed surgical castration for prostate cancer therapy in 1941 (1). Since then, androgen deprivation therapy (ADT) still plays an important role for advanced prostate cancer or prostate cancer patients in elderly patients Due to the recent development of gonadotropin releasing hormone (GnRH) medication, ADT has shifted from surgical castration to medical castration. Leuprorelin was firstly developed in 1985, since then depo medication has come to clinical use in 1989. Goserelin acetate was firstly introduced in 1987 and 3 months depo was introduce in 1999 (2, 3). These GnRH agonist was used as ADT for prostate cancer for four decades. In 2012, degarelix as GnRH antagonist came to clinical use in Japan and 3 months doseage of degarelix was introduce in 2019 (4). Though some differences including the difference of dose persistence, needle size, GnRH agonist or antagonist, either drugs control serum testosterone level (5). On the other hand, the diameter of needle, the duration of efficacy in each drug, and side effect including pain and skin reaction varies in each GnRH drugs. This study examined the patients’ preference of GnRH agonist and antagonist and the difference of taking time for finishing injection of these medicine in the hospital.

A total of 851 hormonal injections were performed from August 2018 to February 2019 in Yokohama City University Medical Center (Yokohama, Japan). None of the patients received generic drugs. Nine cases (1.0%) were received leuprorelin 3.75mg (Leuprin®□), 232 cases (27.3%) were received leuprorelin 11.25mg (Leuprorin SR®□), 187 cases (22.0%) were received leuprorelin 22.5mg (Leuprin PRO®□), 206 cases (24.2%) were receive goserelin acetate 10.8mg (Zoladex LA®□), and 217 cases (25.5%) were received degarelix acetate 80mg (Gonax®□). Institutional review board approved this study (B200800065). All injections were performed by medical nurses subcutaneously after prescription from urologists. The time from ordering hormonal therapy to the time to finish injection were analyzed. The total time per year was calculated each time multiplied by the numbers per year. We also asked a total of 50 patients about the mental and physical load to hormonal injection. The load was evaluated the cost as a following question: ‘‘If the hormoranl injection is able to be change to oral tablet forms, how much cost (Japanese Yen: JPY) will you pay per one injection?’’. When patient who preferred injection to tablet or did not want to change the forms of hormonal therapy, the cost was defined as 0 JPY. The total cost per year was calculated one time cost multiplied by the numbers per year. All patients were received the same GnRH drugs more than two times before this study.

Median (mean +/- SD) time from ordering hormonal therapy to the time to finish injection were 7 (8.9+/-1.7) in leuprorelin 3.75mg, 7 (7.7+/-4.9) in leuprorelin 11.25mg, 8 (8.0 +/- 5.5) in leuprorelin 22.5mg, 6 (7.251+/-3.9) in goserelin acetate 10.8mg, and 10 (11.9+/-6.8) in degarelix acetate 80mg. Time from ordering hormonal therapy to the time to finish injection in degarelix were significantly higher than the rest GnRH hormonal treatments (p<0.001) [Fig. 1a&b].

**Figure 1.**
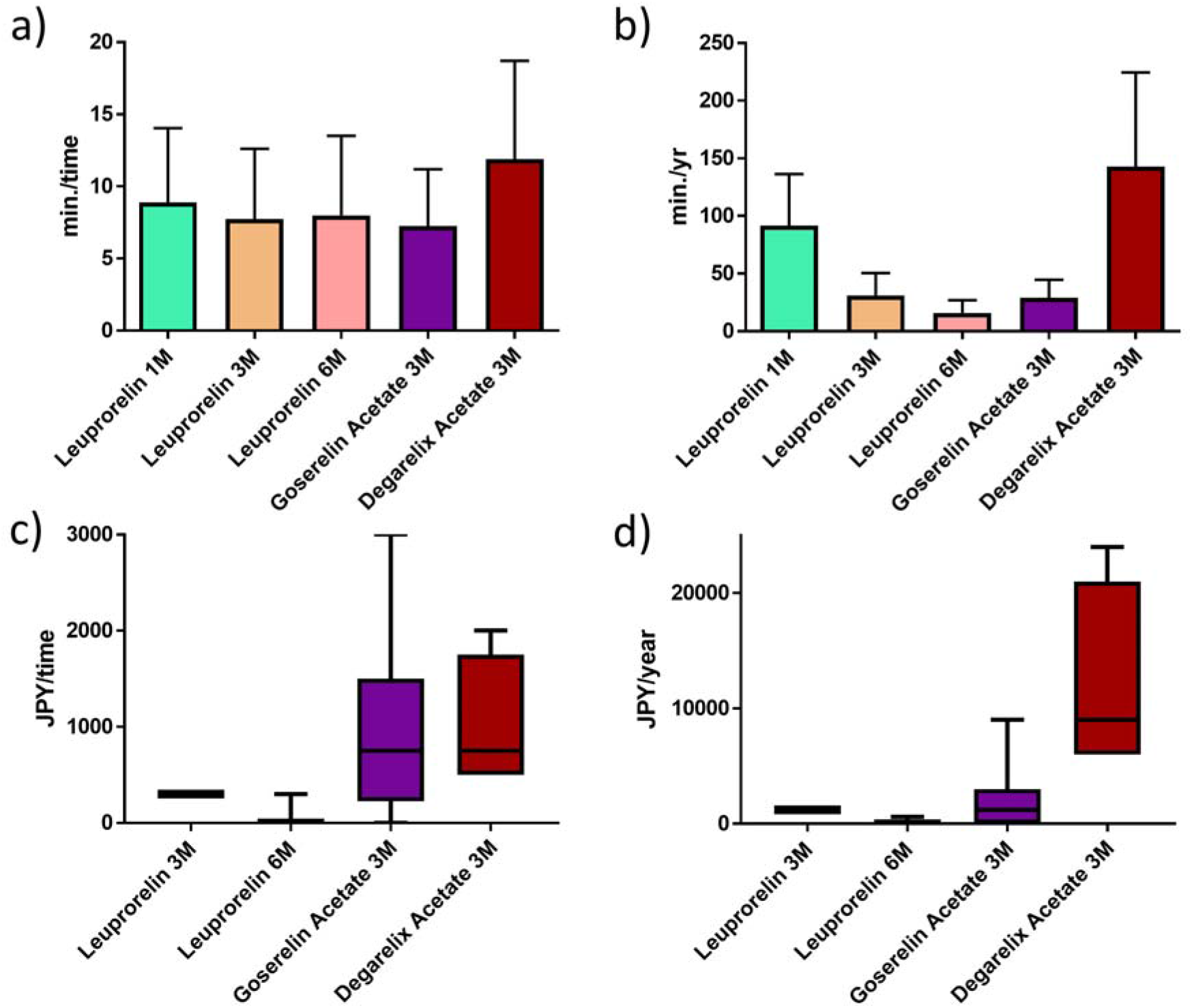
(a) Time from prescription to finish injection in one time. (b) Time from prescription to finish injection in a year. (c) Load of physical and mental costs in each injection. (d) Load of physical and mental costs in a year.

Median (mean +/- SD) cost of load to injection were 300JPY (325.0 +/- 35.4) in leuprorelin 11.25mg, 0.0JPY (40.0 +/- 105.6) in leuprorelin 22.5mg, 750.0JPY (966.7 +/- 1070.8) in goserelin acetate 10.8mg, and 750.0JPY (1000.0 +/- 707.1) in degarelix acetate 80mg. In leuprorelin 22.5mg, 13 of 15 (86.7%) cases did not want to change for oral tablet [Fig.3a&b].

In this study, leuprorelin 22.5mg was able to reduce hospital stays the most, partly because of its 6-month formulation and partly because of its kit formulation. This was due to the fact that leuprorelin and goserelin acetate are in kit form, while degarelix requires dilution of the drug in each case, which is thought to slow down the administration process. In addition, because degarelix can cause skin reactions such as redness and pain, our clinic cools the injection site before injecting it to reduce these skin reaction, which is thought to take more time. With regard to patient burden from the injectable formulation, leuprorelin 22.5mg had the lowest, with little difference in burden from the oral formulation, especially with regard to the 6-month formulation.

This study has the limitation that leuprorelin was injected subcutaneously based on the approval method of administration in Japan, but leuprorelin is injected intramuscular in most of the other county and leuprorelin is injected every three months. Second, since 2019 degarelix acetate 240mg x2injections was introduced as three monthds duration methods in Japan. Thus further investigation is needed (6).

In conclusion, the 6-month formulation of leuprorelin 22.5mg had the lowest time spent in the hospital, and degarelix acetate tended to have the highest patient burden from injections, including pain.

## Data Availability

All data produced in the present study are available upon reasonable request to the authors

## Availability of data and materials

The datasets used and/or analyzed during the current study are available from the corresponding author upon request

## Declarations

### Ethics approval and consent to participate

The present study was approved by the Institutional Review Board of Yokohama City University Medical Center [B200800065]. This was a retrospective study, and additional informed consent was not required by the IRB.

### Consent for publication

Not applicable, in this study.

### Availability of data and materials

Due to ethical restrictions, the raw data underlying this paper are available upon request to the corresponding author.

### Authors’ Contributions

Conceived and designed the experiments: TK, HU. Analyzed data: TK Performed the experiments: TK, HU. Wrote the paper: TK. All authors read and approved the final manuscript.

## Acknowledgements

N/A

